# Enablers and barriers to implementing a digital logistics system: A qualitative evaluation of OpenLMIS for Nigeria’s vaccine supply chain

**DOI:** 10.1101/2025.11.19.25340613

**Authors:** Ifeoluwa Adebola Noiki, Lola Aladesanmi, Suruchi Gupta, Patricia Mechael, Erica Layer, Smisha Agarwal, Dustin Gibson

## Abstract

**Background:** Logistics management information systems (LMIS) strengthen health commodity distribution and decision-making. Nigeria introduced OpenLMIS, an electronic LMIS, in August 2021 to track COVID-19 vaccines, and a routine immunization (RI) module was deployed nationwide in 2023. This evaluation examined enablers and barriers to effective implementation and sustained use.

**Methods:** A qualitative design combined desk reviews of LMIS documents, 42 key informant interviews, and 16 structured observational assessments with Cold Chain Officers. Data collection occurred in three states, Federal Capital Territory, Lagos, and Niger, and nine Local Government Areas (LGA; three per state), which were purposively selected to capture diverse operational contexts.

**Results:** OpenLMIS has been rolled out across all 835 Nigerian cold stores at zonal, state, and LGA levels. Adoption was driven by strong government leadership, particularly the National Primary Health Care Development Agency (NPHCDA), and consistent financial and technical support from partners including Gavi, the Vaccine Alliance, the Clinton Health Access Initiative (CHAI), and UNICEF. Persisting challenges include high staff turnover disrupting continuity; limited user training, often online-only and insufficient where connectivity is poor; unreliable power and internet hindering timely data entry and access; and inadequate, unsustained funding for data bundles, which shifts costs to users and undermines utilization and data quality.

**Conclusion:** OpenLMIS is advancing digitization and visibility of Nigeria’s immunization supply chain for informed decision making. To maximize its benefits, stakeholders should address individual-level disincentives (e.g., turnover, training gaps), invest in reliable power and connectivity, and ensure sustainable financing for data and system operations alongside continued improvement in functionality.

## 1. Introduction

Logistics Management Information Systems (LMIS) are foundational for effective supply chain decisions within public health, managing critical data points such as stock levels, consumption rates, instances of losses, and detailed shipment information. Beyond mere record-keeping, their core functions extend to sophisticated forecasting, precise inventory and distribution planning, comprehensive reporting, efficient order fulfillment, and continuous performance monitoring [1]. Electronic LMIS (eLMIS) significantly enhances these capabilities, primarily by offering more timely data, improved data visibility across the supply chain and facilitating seamless integration with electronic medical records (EMRs)[2,3]. Despite these advantages, many low- and middle-income countries (LMICs) continue to rely on paper-based systems, which often result in data quality issues and delays. While eLMIS facilitates real-time inventory tracking, streamlined workflows, and data-informed decision-making through digitalization, their impact in many LMICs is frequently hindered by fragmented systems and unreliable infrastructure [4]. Prior to the implementation of OpenLMIS –an open-source, cloud-based eLMIS – Nigeria utilised various systems, including Microsoft Navision [5], which was discontinued due to financial and technical challenges, and an Open Data Kit (ODK)-based LMIS [6] that served as an interim solution.

In 2021, the National Primary Health Care Development Agency (NPHCDA) and its partners strategically introduced the Open Logistics Management Information System (OpenLMIS) in Nigeria. This adoption was aligned with the Gavi Target Software Standards (TSS), underscoring a commitment to global best practices in health supply chain management. The primary objectives of this strategic move were to significantly enhance the efficiency of vaccine tracking, provide real-time visibility across the entire supply chain, and harmonize Nigeria’s existing LMIS infrastructure with established international standards [7]. The OpenLMIS’s efficacy was first demonstrated through a successful pilot for COVID-19 vaccine management, conducted at both national and state levels in August 2021. The positive outcomes of this initial implementation paved the way for the development and widespread deployment of a routine immunization (RI) module in January 2023, extending its capabilities to cover a broader range of vaccine-preventable diseases across the country [8]. This paper presents an analysis of the adoption of OpenLMIS in Nigeria, examining the key enablers and barriers to its effective implementation and sustained use. Through a qualitative assessment involving desk reviews, key informant interviews, and structured observational assessments, we explore the user experience to identify key factors influencing the system’s success. The findings from this evaluation offer critical insights for strengthening digital supply chain management for vaccines in Nigeria and similar settings.

## 2. Methods

### 2.1 Study Design

This was a qualitative study that comprised a desk review of relevant Logistics Management Information System (LMIS) documents (e.g. system function definitions), project implementation updates/reports, key informant interviews (KIIs) with stakeholders, and observational assessments of OpenLMIS in practice. The overarching objective of the study was to explore the utilization, perceived benefits, and encountered challenges of OpenLMIS within the context of routine immunization programs and COVID-19 vaccination delivery in Nigeria. The study was designed to elicit insights into user experiences and gather information regarding system functionality to inform future enhancements and improvements.

### 2.2 Ethics

Ethical approval for this study was granted by the National Health Research Ethics Committee (NHREC) in Nigeria, with approval number NHREC/01/01/2007-27/03/2024 and by Johns Hopkins Bloomberg School of Public Health (IRB00029552). Prior to participation in any interviews or observations, all individuals provided informed consent, documented in written form. It was explicitly communicated and thoroughly emphasized that their involvement in the study was entirely voluntary, ensuring no coercion or undue influence.

Participant recruitment spanned the period of 25^th^ June 2024 to 12^th^ July 2024.

### 2.3 Sampling and Sample Size

The study was conducted across three purposively selected states: the Federal Capital Territory (FCT), Niger, and Lagos. These locations reflect a level of geographical diversity, variations in existing infrastructure, and differing levels of OpenLMIS utilization (low, medium and high), thereby ensuring a comprehensive understanding of the system’s performance in varied contexts. Within each of these states, the research included visits to the zonal cold store, the state cold store, and three distinct LGA-level cold stores. A total of 42 in-depth key informant interviews (KII) were conducted with a range of stakeholders (program managers and cold chain officers) at the national, zonal, state, and LGA levels, capturing perspectives from across the entire supply chain hierarchy, except at the health facility level where OpenLMIS is not currently deployed. Complementing these KIIs, 16 systematic observational assessments of the Cold Chain Officers were carried out at Zonal, State and LGA cold stores. These observations were critical for gaining practical insights into the platform’s actual usage, identifying workflow challenges, and directly assessing users’ proficiency in navigating and operating OpenLMIS in real-world settings.

Participants were identified and recruited through purposive sampling in collaboration with national and state-level immunization program managers. Respondents were selected based on their direct involvement with the OpenLMIS platform and included Cold Chain Officers, Immunization Program Managers, and LMIS focal persons across all tiers of the immunization supply chain (National, Zonal, State and LGA). This approach ensured a range of perspectives from individuals in various roles, enhancing the relevance and applicability of the study’s findings.

### 2.5 Theory of Change and Tools Development

Our study was guided by a Theory of Change (ToC, see S1 Annex) that outlines how OpenLMIS is expected to strengthen vaccine logistics and, ultimately, immunization outcomes. The ToC informed the development of our study objectives and the design of our data collection tools, semi-structured interview guides and an observational tool. These instruments were designed to test the ToC’s underlying assumptions by capturing stakeholder experiences and assessing user digital literacy. Interview questions examined system adoption and use, data flows and quality, decision-making processes, and perceived changes in stock visibility and vaccine delivery. The ToC also defined thematic domains for the interview guides and analytic framework, including enablers, barriers, user experience, functionality, and integration.

### 2.6 Data Collection Process

Experienced and trained research assistants (58% male; 42% female) conducted interviews using the semi-structured interview guides (See S2 Annex and S3 Annex). We conducted KIIs with national and sub-national program managers, cold-chain officers, and implementing partners. Data collection continued until thematic saturation was reached across three purposively selected states (Federal Capital Territory, Niger, and Lagos) and nine LGAs. Most interviews were conducted face-to-face, in English, and lasted 45–60 minutes. Interviews were audio-recorded for verification and quality checks. Observational assessments used a structured checklist (See S4 Annex) to assess users’ ability to perform core OpenLMIS tasks, including: creating physical inventories, issuing orders, confirming shipments, making adjustments, transferring stock between programs, generate stock-balance/expiry/distribution reports, and managing cold chain equipment. Interviewers also documented user feedback, challenges, and contextual factors observed during platform use. Prior to fieldwork, courtesy visits introduced the study to sites and addressed potential administrative barriers. Interviewers had no prior engagement with participants.

### 2.7 Data Analysis and Coding Process

Codes were developed deductively and inductively. Thematic categories focused on enablers, barriers, user experience, system functionalities, and suggestions for improvement. All interviews and field notes were transcribed then subjected to a stepwise thematic analysis process: 1) comprehensive reading for context; 2) open coding to label salient segments; 3) axial coding to cluster related codes and map relationships; and 4) selective coding to integrate core categories into themes aligned with research questions [9]. Observational assessments were included in coding for user-related factors, challenges and recommendations for the use of the OpenLMIS. Also, items on the checklist used for the observational assessment were assigned scores, and an overall score was computed based on the maximum available score for each individual, and category. Credibility of findings was strengthened through triangulation of KIIs, observations, and documents (identifying points of divergence and complementarity to further contextualize findings); enumerator debriefings (consensus meetings) to minimize bias and validate emerging interpretations; and stakeholder validation with select respondents to confirm the accuracy and relevance of emergent findings.

## 3. Results

### 3.1 Demographic Characteristics of the sample

We conducted a total of 42 KIIs and 16 observational surveys (see Table 1). KIIs were 63% (n=26) male and 37% (n=16) female (See Table 1). By role, 43% (n=18) were cold chain officers and 57% (n=24) were program managers overseeing immunization supply chain functions. The distribution of KIIs across administrative levels was as follows: 38% (n=16) were at the LGA level, 31% (n=13) at the State level, 10% (n=4) at the Zonal level, and 21% (n=9) at the National level. The distribution of KIIs by administrative level, stakeholder role, and organization for the three states are presented in Table 2.

**Table 1.** Demographic Characteristics of Key Informant Interview (n=42) and Observational Assessment Participants (n=16)

|  | Key Informant<br>Interviews | Observational<br>Assessment |
| --- | --- | --- |
| Demographic | n (%) | n (%) |
| **Gender** |  |  |
| Male | 26 (63) | 11 (69) |
| Female | 16 (37) | 5 (31) |
| **Stakeholder Role** |  |  |
| Program Manager | 24 (57) | 2 (13) |
| Cold Chain Officer | 18 (43) | 14 (88) |
| **Administrative level** |  |  |
| National | 9 (21) | 2 (13) |
| Zonal | 4 (10) | 2 (13) |
| State | 13 (31) | 4 (25) |
| Local Government Area (LGA) | 16 (38) | 8 (50) |

**Table 2.**
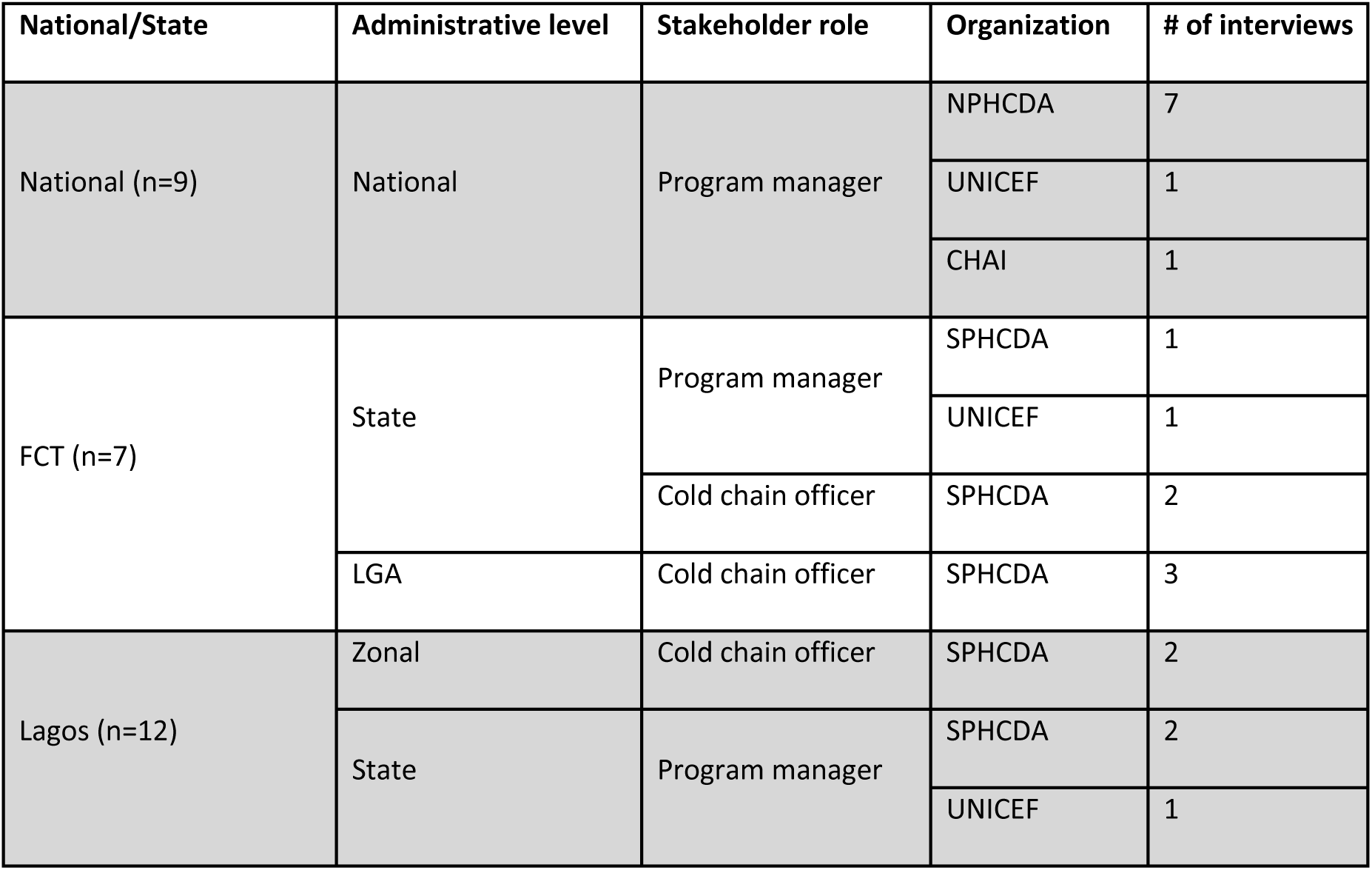
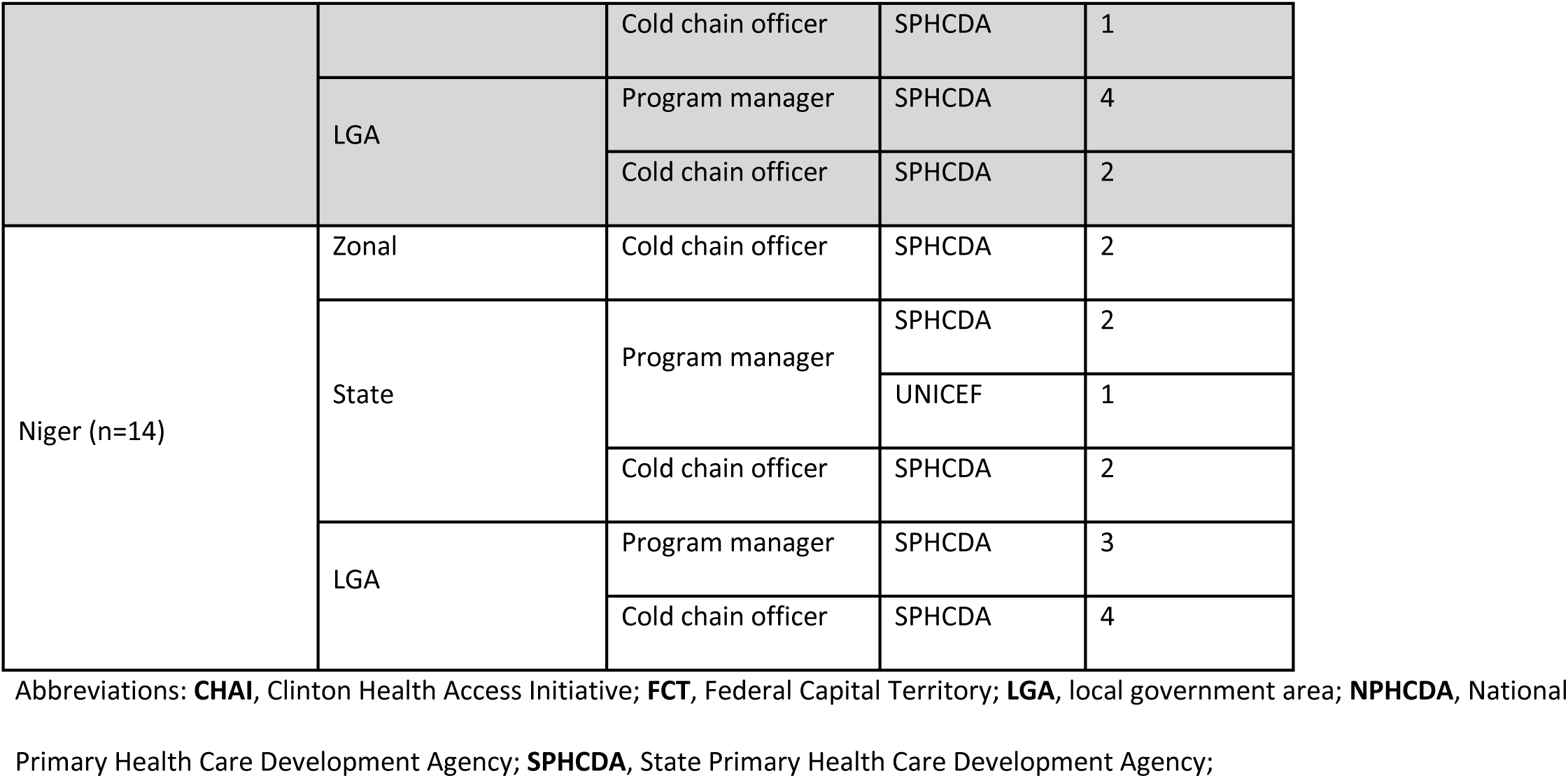
Distribution of Key Informant Interview Participants by Administrative Level, Stakeholder Role, and Organization.

### 3.2 Perceived benefits of OpenLMIS

OpenLMIS has been successfully rolled out across all 835 Nigerian cold stores, encompassing Zonal, State, and LGA levels. The integration of OpenLMIS at higher levels of the supply chain has resulted in enhanced efficiency, a shift in logistics personnel roles, and more proactive supply Chain Management. This evolution underscores the escalating importance of digital tools and data-driven strategies in logistics, promoting enhanced accountability, transparency, and necessitating continuous skill development for a workforce more adept at leveraging technology for improved public health outcomes.

*“The use of OpenLMIS has led to streamlined processes and reduced paperwork, allowing staff to focus more on vaccination utilization. It has also enhanced accountability, better tracking of stock and transparency. Some staff roles have also shifted to include more data management and analysis tasks”- State Program manager, SPHCDA*

The efficiency brought about by OpenLMIS allows for a more proactive approach to supply chain management. By providing real-time data and insights, the system enables stakeholders to identify and mitigate potential issues before they escalate into larger crises. This foresight ensures a more reliable and responsive system for critical vaccine distribution, minimizing stockouts, optimizing cold chain integrity, and ultimately improving public health outcomes by ensuring vaccines reach their intended recipients effectively and consistently.

*“OpenLMIS enhances the management of vaccine supply chains by providing real-time data on vaccine stock levels, distribution, and usage. This improved data visibility allows us workers and our senior management to make informed decisions about vaccine utilization across all levels within the state.” - State Program manager, SPHCDA*

### 3.3 Perceived Enablers and Barriers to OpenLMIS adoption and use

The study identified several perceived factors that influenced the adoption and use of OpenLMIS in Nigeria. These factors, grouped into organizational/environmental, user-related, and technological themes, encompass the different enablers and barriers that shape the environment for the implementation and sustainability of the OpenLMIS across Nigeria (See Table 3).

**Table 3.** Summary of perceived enablers and barriers to OpenLMIS adoption by theme.

| Theme | Factor | Examples |
| --- | --- | --- |
| Organizational/<br>Environmental | Government commitment and partner support **(E)** | Strong leadership from NPHCDA and consistent support from Gavi, CHAI, and UNICEF partners |
|  | Financial constraints **(B)** | Lack of sustainable funding for internet data forces users to pay out-of-pocket |
|  | Inadequate training **(B)** | Often inconsistent, insufficient, or delivered virtually with poor connectivity |
|  | Insufficient local user-support **(B)** | Error corrections requires national remediation leading to delays and disruptions |
|  | Infrastructure and equipment gaps **(B)** | Unreliable internet connectivity and power supply with no offline mode availability |
| User-related | Financial disincentives **(B)** | Staff often fund internet data from their own salaries thus reducing motivation to use |
|  | Human resource management **(B)** | High staff turnover, often due to political appointments, disrupts knowledge continuity |
|  | Increased workload **(B)** | At the LGA, requirements for dual data entry (paper and OpenLMIS) seen as burdensome as inefficient |
|  | Digital literacy and platform proficiency **(E)** | High proficiency in using platform by cold chain officers |
| Technological | Highly valued platform features **(E)** | Users praised features like automated reminders for vaccine expiration and data validation checks |
|  | Platform features needing improvement **(B)** | Users reported slow system speed, a cumbersome user interface, and system errors like generation of duplicate IDs |
|  | Requested platform features **(B)** | Key features requested include offline client, barcode scanning (in bulk) and interoperability with other systems |

Abbreviations: **(B)**, Barrier; **(E)**, Enabler; **CHAI**, Clinton Health Access Initiative; **LGA**, local government area; **NPHCDA**, National Primary Health Care Development Agency;

#### 3.3.1 Organizational/ Environmental factors

##### Government commitment and partner support

The success of OpenLMIS’s implementation was perceived to be due to the strong commitment and collaborative efforts of various stakeholders. These included key implementing partners such as CHAI, Gavi, and UNICEF, as well as government bodies, notably the National Primary Health Care Development Agency (NPHCDA) and its National Logistics Working Group (NLWG). This support was critical for both the initial rollout and the sustained utilization of the OpenLMIS system across Nigeria.

*“Technical support from the partners and NPHCDA has been very crucial”- State Program manager, SPHCDA*

Gavi, CHAI, and UNICEF provided multifaceted support. This encompassed the initial onboarding training and periodic refresher training sessions, which were facilitated by CHAI and the NPHCDA. Furthermore, UNICEF offered ongoing technical support, primarily delivered through convenient and accessible channels such as WhatsApp and Zoom. This support helped users resolve operational issues and maximize the full functionality of the system, facilitating usage. A notable observation was that higher levels of engagement with OpenLMIS were consistently observed in states and LGAs that demonstrated strong leadership and maintained consistent follow-up protocols.

##### Financial constraints

Financial constraints were identified as a critical challenge, primarily due to the lack of sustainable funding for internet data, which poses ongoing difficulties for OpenLMIS implementation and usage. While external support from CHAI and UNICEF provided temporary relief, the absence of a long-term financial plan leaves the system vulnerable to disruptions.

*“When the roll-out came; there was no support for data provision and OpenLMIS is data intensive; CHAI however provided support for this at the LGA level and also provided training and re-training. CHAI also provides virtual support and training. However, the major issues are now as regards the provision of internet services.” - State Program manager, SPHCDA*

This absence of dedicated funding also affects the perceived value of OpenLMIS, as resource allocation from government offices to support its implementation has been inadequate. There is a perceived need for increased government ownership of the OpenLMIS implementation process to ensure its sustainability and maximize its benefits. This includes establishing consistent budget lines for internet connectivity, maintenance, and ongoing training to prevent service interruptions and ensure the system’s longevity.

##### Inadequate training

The effectiveness of OpenLMIS capacity building activities provided were hampered by inconsistencies in both the frequency and quality of sessions. Training programs were often irregular and, at times, demonstrably insufficient, underscoring a pressing need for more consistent and comprehensive educational initiatives.

*“Capacity building was a real issue for adoption; it took a lot of training and retraining to understand how the system works.”- State Program manager, SPHCDA*

This was particularly critical for new users or individuals who had missed previous sessions, as their lack of initial exposure significantly impeded their ability to fully leverage the system. This challenge was acutely observed in states with low OpenLMIS utilization, where participants frequently cited incomplete training or recent employment as primary reasons for their limited proficiency across various system functionalities. The initial rollout for COVID-19 vaccination, for instance, relied heavily on virtual training modules. However, these were largely met with resistance and disinterest from some stakeholders, primarily due to existing digital literacy gaps and significant participation issues stemming from unreliable internet access, especially prevalent at the state and LGA levels.

##### Inadequate local user-support

Another significant impediment to the system’s effectiveness is the lack of localized technical support, particularly for correcting errors that arise from user input or system malfunctions. Currently, many system errors, such as duplicate transaction IDs or discrepancies between system data and downloaded reports, require intervention from national-level personnel. This centralized troubleshooting method causes substantial delays, interrupts user workflows, and reduces overall efficiency.

*“If I make a mistake I can’t correct it and that mistake is what will be seen on the OpenLMIS. The OpenLMIS desk officer in Abuja should try from time to time to be able to make corrections since we can’t delete our mistakes by ourselves.” - Zonal Cold Chain Officer, SPHCDA*

The absence of readily available local technical assistance means that issues that could be resolved at a lower administrative level are instead escalated, negatively affecting system performance and user morale. This reliance on a central authority for even minor technical issues creates a bottleneck, leading to frustration among users and a slower resolution time for critical problems.

##### Infrastructural and equipment gaps

Infrastructural limitations were identified as a critical impediment to the effective implementation and sustained use of OpenLMIS. A primary concern is the unreliable internet connectivity prevalent across many regions of Nigeria. This issue is exacerbated by the current non-utilization of the OpenLMIS offline client, which renders users entirely dependent on consistent internet access—a resource that is frequently unavailable or unstable, particularly in remote areas and at lower levels of the supply chain. The resulting persistent network downtime disrupts operational workflows, leading to delays in task completion and subsequently discouraging consistent system adoption. Beyond connectivity, other critical infrastructural deficiencies compound these challenges, notably an inconsistent power supply. In regions afflicted by unreliable electricity, users encounter substantial difficulties in maintaining the charge of their computing devices, further curtailing their capacity to engage with the system consistently. Furthermore, frequent concerns were raised regarding the availability, maintenance, and security of computing equipment. Reports of damaged or stolen devices directly underscore how these issues impact the accessibility and sustained use of OpenLMIS in specific locations. These widespread infrastructural challenges collectively contribute to a less than optimal environment for the full realization of OpenLMIS’s potential.

#### 3.4.2 User-related factors

***Financial Disincentives:*** A primary barrier to user motivation stems from prohibitive data costs. Personnel, especially at the LGA level, often fund internet services out-of-pocket due to a lack of dedicated budgetary allocations.

*“Out of pocket funding of data has been a huge challenge; with delays in paying salaries, which is not unique to just the supply chain but to service delivery in general.” - National Program manager, NPHCDA*

This issue is exacerbated by broader financial constraints, including delayed salaries and limited operational budgets, which collectively reduce staff commitment and motivation. The absence of financial incentives, such as performance-based bonuses or allowances for data usage, further demotivates staff from consistently engaging with the OpenLMIS system.

##### Human Resource Management

Frequent turnover of personnel, largely driven by political leadership/governance and influence in human resource decisions, significantly disrupts the continuity and institutionalization of OpenLMIS knowledge.

*“Change in governance often affects the appointment of the cold chain officers… creating a huge gap in knowledge and competency.”- National Program manager, NPHCDA*

Newly appointed staff members often lack the necessary skills and receive insufficient or no training, leaving them unprepared to effectively use the system. This lack of adequate human resources, both in terms of quantity and quality, poses a significant barrier to the sustainable adoption and effective utilization of OpenLMIS. The constant need to onboard and train new staff diverts resources and time, hindering progress and potentially leading to errors or underutilization of the system’s capabilities. Furthermore, the absence of a stable and skilled workforce undermines efforts to build internal capacity and create a resilient system that can adapt to evolving needs and challenges.

##### Increased Workload

At the LGA level, the requirement to maintain parallel paper-based and electronic records creates a substantial increase in workload. This dual data entry is perceived as burdensome, as the cold chain officers need to collate paper-based records from several health facility stores and enter them manually into the LGA store records and then into OpenLMIS.

*“It has added to our problem because we don’t have supporting hands. We enter the ledger and the OpenLMIS at the same time, so OpenLMIS adds to our workload”- LGA Cold chain officer, SPHCDA*

This duplication leads to inefficiencies and, in some cases, data entry errors, particularly when handling large volumes of data or multiple tasks. The time-consuming nature of this process diverts staff from other critical duties, potentially impacting overall supply chain efficiency and accuracy. In contrast, at higher levels of the supply chain (State, Zonal, and National), OpenLMIS has been perceived to positively impact workflows by significantly reducing reliance on manual paperwork. This digitalization has streamlined logistics processes, allowing staff to reallocate their focus to more critical tasks such as vaccine data analysis and utilization for informed decision-making.

##### Digital literacy and platform proficiency

Findings from observational assessments indicate a generally high level of OpenLMIS knowledge, proficiency, and digital tool literacy among cold chain officers. The aggregated performance data indicates that, on average, they were able to successfully complete 85% of the tested tasks without encountering significant challenges (See Table 4).

**Table 4.** Observational assessment of cold chain officer’s ability to complete OpenLMIS task.

| OpenLMIS task | n (%) |
| --- | --- |
| Create physical inventory | 15 (94) |
| Issue an order | 15 (94) |
| Confirm shipment | 13 (81) |
| Make stock adjustments | 16 (100) |
| Transfer stock from one program to another | 14 (88) |
| Generate stock balance report | 16 (100) |
| Generate report showing stock expiry status | 11 (69) |
| Generate stock distribution report | 12 (75) |
| Manage cold chain equipment | 11 (69) |
| **Total** | **85.4%** |

#### 3.3.1 Challenges with Specific OpenLMIS Tasks/Functions

Despite the overall high proficiency, certain functionalities within OpenLMIS consistently presented difficulties for users, suggesting areas ripe for targeted improvement. In creating physical inventory, users frequently described this process as cumbersome, particularly when handling bulk submissions of inventory data. They suggested improvements such as allowing individual product updates, which would provide more flexibility, and the ability to hide out-of-stock or expired batches directly within the OpenLMIS interface to reduce visual clutter and potential for error.

*“I should be able to update products individually” - State Cold Chain Officer, SPHCDA*

*“I’d prefer to be able to hide every batch that is zero, to make the interface less cumbersome” - National Program manager, NPHCDA*

For issuing orders, users reported challenges related to accurate product listing and, notably, the lack of system functionality to withdraw or correct previously issued products. This rigidity led to recommendations for system enhancements, including features like auto-summation of vaccine quantities to prevent manual calculation errors and better management of product expirations to ensure only viable stock is issued.

*“We need to upgrade the system in such a way that, when we issue out to a facility and discover they don’t use it we should be able to withdraw it and issue it out to another facility that need it urgently.” - LGA Cold Chain Officer, SPHCDA*

For managing Cold Chain Equipment (CCE), users reported that this functionality was a significant challenge, largely attributed to insufficient training provided to users and the inherent complexity of the system’s interface for this specific task. Users expressed a strong need for more comprehensive training modules dedicated to CCE management and fundamental system improvements to simplify the process.

For confirming shipments, some users advocated for improved external communication protocols (e.g., automated notifications when shipments are dispatched) and enhanced record-keeping capabilities within OpenLMIS to facilitate easier reconciliation and verification of incoming deliveries.

*“If I have 200 antigens and want to make 210, when I click on the antigen there should be a notification that will show my stock.” - LGA Cold Chain Officer, SPHCDA*

*“It will be better if we can get notifications for low stock levels or upcoming expirations via SMS or email to serve as the reminder” - LGA Program manager, SPHCDA*

Conversely, “generating stock reports” (encompassing balance, expiry, and distribution reports), “rectifying stock balance,” and “transferring stock items” were generally found to be relatively easy for users to perform. Exceptions occurred primarily in cases where prior training was clearly lacking, underscoring the importance of adequate instruction across all functionalities.

#### 3.4.3 Technological factors

##### Highly-valued platform features

The most appreciated functions of OpenLMIS included reminders and data validation capabilities. Reminder and alert functions were highly valued by users as it significantly aided in monitoring vaccine expiration dates, preventing stockouts by prompting reorders, and ensuring accurate physical inventory counts through timely notifications for reconciliation. However, a recurring point of feedback was the inconsistency of these alerts, with some users reporting delayed or missed notifications. In regards to data validation, the system’s ability to detect and highlight data entry errors, such as incorrect vaccine counts, incorrect unit of measure, or discrepancies between expected and actual inventory levels, was also a frequently praised feature. This functionality helped maintain data integrity and reduce manual reconciliation efforts.

##### Platform features needing improvement

According to users, OpenLMIS needs significant improvement across several key areas to enhance its effectiveness and user satisfaction. A primary concern among users is the overall system performance, specifically the reported slow system speed, which hinders efficient operations. This is likely linked to poor internet connectivity and requests for offline capabilities were requested.

*“If we can be using it offline so we can use it even where there is no network connection…because without data, you cannot open it and its really frustrating at times when loading”- Cold chain officer*

Furthermore, issues with browser compatibility frequently arise, leading to an inconsistent user experience. The user interface (UI) is often described as cumbersome and overly complex, making navigation and data entry challenging. A recurring pain point is the difficulty users encounter when attempting to update stock records, a critical function for inventory management.

Users encounter various system errors that compromise data integrity, such as the generation of duplicate transaction IDs, which can lead to confusion and inaccuracies. Discrepancies between data displayed within the system and information presented in downloaded reports have also been noted, undermining trust in the system’s accuracy. Challenges persist in extracting comprehensive summary reports, limiting the ability to gain quick insights into logistics operations. Accessing historical transaction receipts for auditing and reconciliation purposes has proven difficult, as has the efficient archiving of data for long-term trend analysis and strategic planning. Lastly, the integration and recording of Cold Chain Equipment (CCE) data present significant difficulties for users. The underutilization of this crucial feature is often attributed to inadequate training provided to users on its functionalities and a general lack of user-friendliness within the CCE module, deterring its effective adoption and use.

##### Requested platform features

Several OpenLMIS functions (yet to be configured into Nigeria’s OpenLMIS) were identified as crucial for improving its effectiveness, including: barcode scanning (preferably at the carton level for high volumes) to enhance stock-taking efficiency and expedite data entry; temperature monitoring to ensure vaccine integrity; offline client to decrease dependence on internet connectivity (as only the web-based version is currently in use); and interoperability to connect logistics data with service delivery data, such as vaccine utilization reports from the National Health Management Information System (NHMIS-DHIS2) and the Electronic Management of Immunization Data (EMID) platform, reducing reliance on manual triangulation via platforms like Thrive360.

## 4. Discussion

This qualitative study of Nigeria’s national OpenLMIS rollout provides critical insights into the complex interplay of factors shaping the adoption and use of digital health technologies in a LMIC setting. The successful deployment of OpenLMIS across all 835 cold stores demonstrates the significant progress made toward strengthening Nigeria’s vaccine supply chain. However, our findings reveal that this successful implementation is moderated by persistent challenges related to workforce capacity, financial ownership, and foundational infrastructure, which threaten the system’s long-term sustainability and reflect broader themes in the global dial health literature[10].

A primary learning from this evaluation is that strong government ownership is a critical enabler for success. The leadership of the NPHCDA, and supported by partners like Gavi and CHAI was perceived to be one of the key drivers of the national rollout. This lesson is similar to findings from a multi-country evaluation which found that full government ownership was essential for the long-term success of digital health tools[11]. In Rwanda, the government’s decision to build its immunization registry on the national DHIS2 platform developed local expertise and ensured sustainability. In contrast, there was an overreliance on external partners for a customized system in Tanzania which led to its discontinuation in many facilities once external support was phased out [11]. The need for local empowerment is further underscored by our own findings, which revealed that the reliance on national-level personnel for troubleshooting and error resolution created significant delays and user frustration which may impact system efficiency.

However, it is important to distinguish between strong programmatic ownership demonstrated by the NPHCDA’s leadership and the still-maturing state of its financial ownership. Our findings show that while the government has strong stewardship over policy and implementation, the long-term operational costs, such as internet data for users, are not yet secured through dedicated domestic budget lines; thereby creating a sustainability paradox where the system is a national priority, yet its daily functioning remains vulnerable to out-of-pocket payments by frontline workers. This threat of donor dependency is further exemplified by experience in Ethiopia, where a successful mobile stock management tool was abandoned after project funding ended and with no exit or transition strategy in place[12]. This situation reflects a common stage in the digital health maturity model, where political will often outpaces the slower process of integrating a system’s recurrent costs into the national budget.

Beyond sustainability, the effectiveness of the system is consistently challenged by foundational human resource and infrastructure gaps. Issues of high staff turnover, inconsistent user training, and the burdensome requirement of dual data entry are not only operational hurdles, but systemic weaknesses. These challenges reflect a well-documented pattern in LMICs, where fragmented health information systems and weak human resource planning strain the health workforce and can lead to burnout, ultimately undermining the utility of digital tools[13]. Similarly, the obstacles of unreliable internet and power remain pervasive barriers to digital health adoption across the continent, limiting the potential of otherwise well-designed systems[14].

This study has several limitations. First, interviews and observational assessments were conducted in three purposively selected states. While selected to represent diverse operational contexts, these sites may not be representative of the experiences across all 36 states in Nigeria, limiting the generalizability of our findings. Second, our data collection relied on key informant interviews which may be subject to social desirability bias. We sought to mitigate this through triangulation with observational data and a review of system documents. Furthermore, our evaluation was qualitative in nature and did not include a secondary analysis of quantitative data from the OpenLMIS system itself, which could have provided objective metrics on system utilization and reporting. Lastly, this study focused exclusively on the use of OpenLMIS for the routine immunization program; the findings may not be applicable to its use for other health commodities or in different program areas.

In conclusion, the national scale-up of OpenLMIS represents a significant achievement in Nigeria’s progress toward a digitally enabled vaccine supply chain. The successful adoption and use of the system, driven by strong government ownership and partner collaboration, provides a model for other large-scale digital health implementations. However, for this success to be sustainable, the persistent challenges, like high staff turnover, inconsistent user proficiency, and infrastructure gaps, must be addressed. Realizing the full potential of OpenLMIS will require a continued investment in both the technology and the people and systems that support it.

## Data Availability

The data supporting the findings of this study are not publicly available due to ethical considerations and restrictions, as the qualitative interview transcripts contain information, coupled with the small sample size, that could easily compromise participant confidentiality. De-identified data may be made available upon reasonable request to the corresponding author, Ifeoluwa Noiki, at

## Acknowledgments

The authors thank all partners and participants for their support.

## Supporting Information

### S1 Annex: Theory of Change: OpenLMIS implementation and use in Nigeria

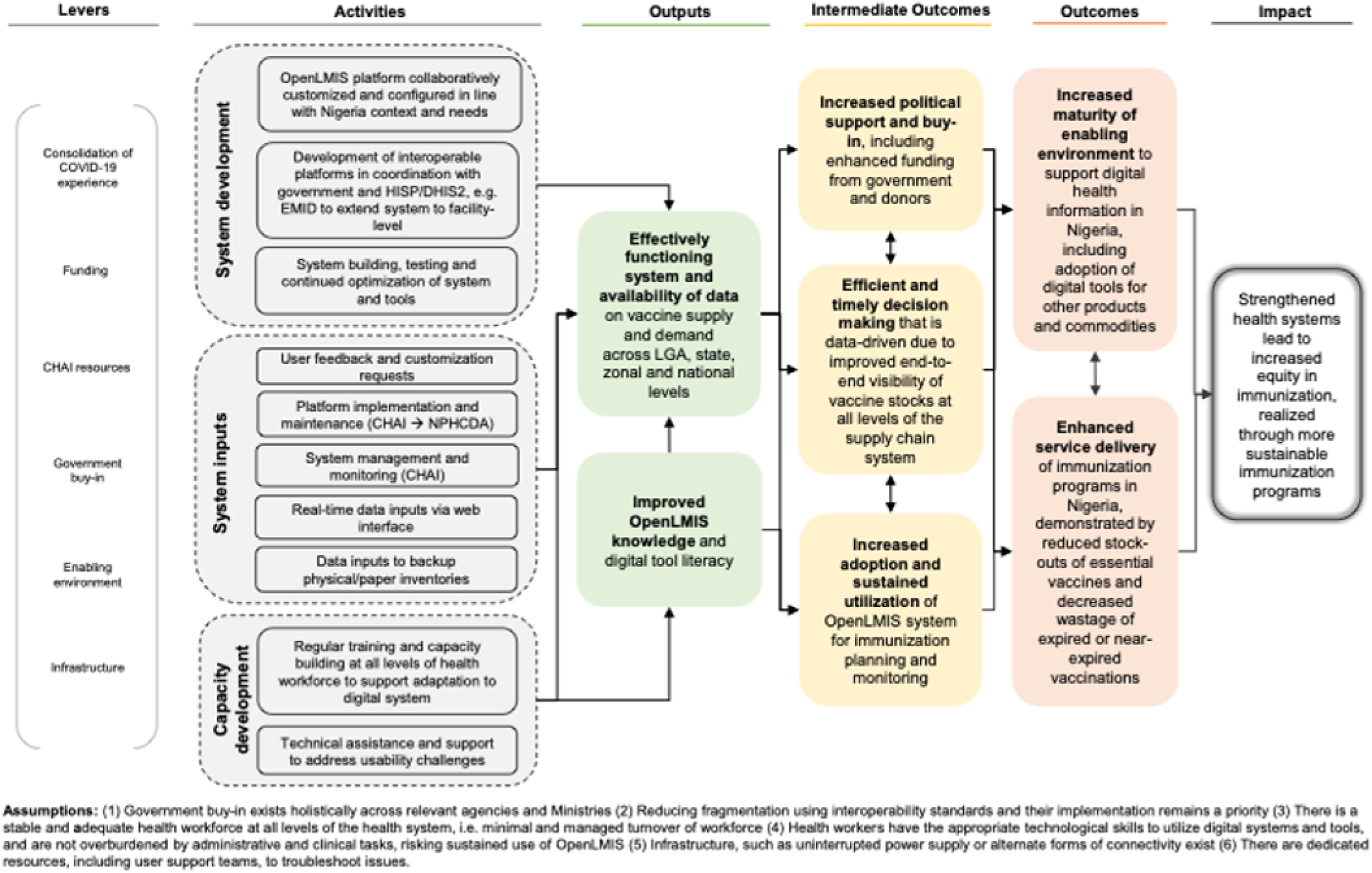

### S2 Annex: Key Informant Interview Guide: Implementers and Program Managers

*CHAI, NPHCDA, Director Logistics and Health Commodities, Members of the National Logistics Working Group. National Strategic Store - Logistics Officers, State Director Immunization/ Disease Control, and Vaccine Security and Logistics Officers - UNICEF*

**Respondent’s Gender**: Male Female

1. **OpenLMIS and its role within the EPI Programme and beyond**

a. What was the rationale for the rollout process of OpenLMIS for COVID-19 vaccination and subsequently Routine Immunization (RI) implementation? *Are there any differences in how the application is used in RI vs COVID-19 vaccination*?
b. Describe the role OpenLMIS currently plays within the EPI program. *How do you use the OpenLMIS on a daily, weekly, or monthly basis?* *To what extent do you rely on OpenLMIS data for planning immunization campaigns, allocating resources, and monitoring program performance? Please give examples of how you used OpenLMIS for any of these functions*.
c. How has OpenLMIS impacted your decision-making and program management?
d. To what extent is the OpenLMIS interoperable with other information systems (e.g.DHIS2)?
e. Is the OpenLMIS already being used in other health areas (i.e. for other health commodities)
2. **Barriers and facilitators to adoption, data quality, and use of OpenLMIS**

a. What are the main barriers to the adoption, use, and data quality within OpenLMIS? *Probe for any specific user groups (Cold Chain officer/Program manager, male or female) within the program/supply chain who have difficulty adopting or using OpenLMIS effectively, and why* *Probe further for specific factors (individual, organizational, and/or technological) Probe for variations in OpenLMIS usage in different States/LGAs and reasons for this*. *Probe for barriers hindering gender equity in access to and utilization of OpenLMIS, and what strategies can be employed to address these barriers effectively?* *Probe for any traditional gender roles and stereotypes within organizational facilities, and how these impacts can be mitigated*
b. What factors have facilitated the successful implementation and use of OpenLMIS? *What strategies have been successful in overcoming barriers to OpenLMIS adoption & improving data quality?* *Which organizations are providing funding to system implementation and maintenance?* *How often are training activities around LMIS conducted (who implements training, what is the frequency?) Do you think there are enough resources directed towards openLMIS?*
3. **Perceptions on the link between OpenLMIS data use and immunization coverage**

a. Have you noted any links between OpenLMIS data use and immunization coverage, completion, and timeliness? *Describe specific correlations between data use and improved program outcomes (e.g., higher coverage rates in higher LMIS usage locations). Is there any evidence to support these findings?*
b. Has the use of the OpenLMIS led to any unexpected effects on vaccine delivery or staff roles? Describe these changes.
4. **Learnings and recommendations from OpenLMIS utilization for COVID-19 vaccination**

a. What unique lessons were learned (positive and negative) from using OpenLMIS for COVID-19 vaccination, and what recommendations do you have based on these lessons that can be applied to future large-scale campaigns and outbreak response in Nigeria or other countries? *What factors facilitated such rapid national scaling of the OpenLMIS during Covid ((infrastructure to support OpenLMIS, policy changes, political buy-in)?* *Are there any specific features or functionalities you would recommend adding to OpenLMIS to prepare for future outbreaks or mass vaccination needs?*

### S3 Annex: Key Informant Interview Guide: Cold Chain Officers

*Zonal Cold Chain Officers, State Cold Chain Officers, Vaccine Security and Logistics Officers - UNICEF, Satellite Cold Chain Officers, LGA Cold Chain Officers*.

**Respondent’s Gender**: Male Female

1. **The stock management system and flow of data from paper-based to OpenLMIS**

a. Describe your role and the stock management system you currently use for vaccine stocks
b. Describe the flow of data from health facilities to higher levels of the supply management system. *How often were there any discrepancies between recorded and actual stock levels? Can you share any specific examples? Is this a major problem?*
2. **OpenLMIS user experience, benefits, enablers, and barriers**

a. What are the main benefits of using OpenLMIS, compared to the paper-based system? *Probe for specific examples of how OpenLMIS has improved their work*.
b. How easy is OpenLMIS to use in your daily work? *Probe for specific tasks in their daily work that OpenLMIS makes easier or harder. Probe for how well OpenLMIS integrates with their existing workflow*. *If it integrates well, probe for an example of how OpenLMIS integrates well with their workflow. If not, probe for where it creates friction*.
c. Are there any features of OpenLMIS you find particularly helpful or unhelpful? *Probe if there are any features or functionalities they wish OpenLMIS had that they need to perform their job effectively, or any existing features they rarely use, and why*. *Probe the value of the decision-support algorithms, alerts, and reminders, and if this actually helped them prevent stockouts or make better decisions. Ask for an example*.
d. What factors have enabled you to effectively use OpenLMIS in your work? *Probe further for specific factors (individual, organizational/environmental, and/or technological)*. *Ask them to describe the training and support they have received to enable the implementation of OpenLMIS. How often are training activities around LMIS conducted (who implements training, what is the frequency?)* *Do you think there are enough resources directed towards openLMIS? Do you think its valued by those in charge? Which organizations are providing funding to system implementation and maintenance?*
e. What challenges have you experienced with using OpenLMIS? *Probe further for specific factors (individual, organizational/environmental, and/or technological)* *Ask if they feel they have the necessary skills and training to use OpenLMIS effectively. If not, ask what additional support they need*.
3. **Recommendations for improvement**

a. What improvements would you suggest to make OpenLMIS more effective for future mass vaccination campaigns as well as routine immunization? *Have there been any improvements/investments into openLMIS since it was launched*

### S4 Annex: OpenLMIS Observation Checklist

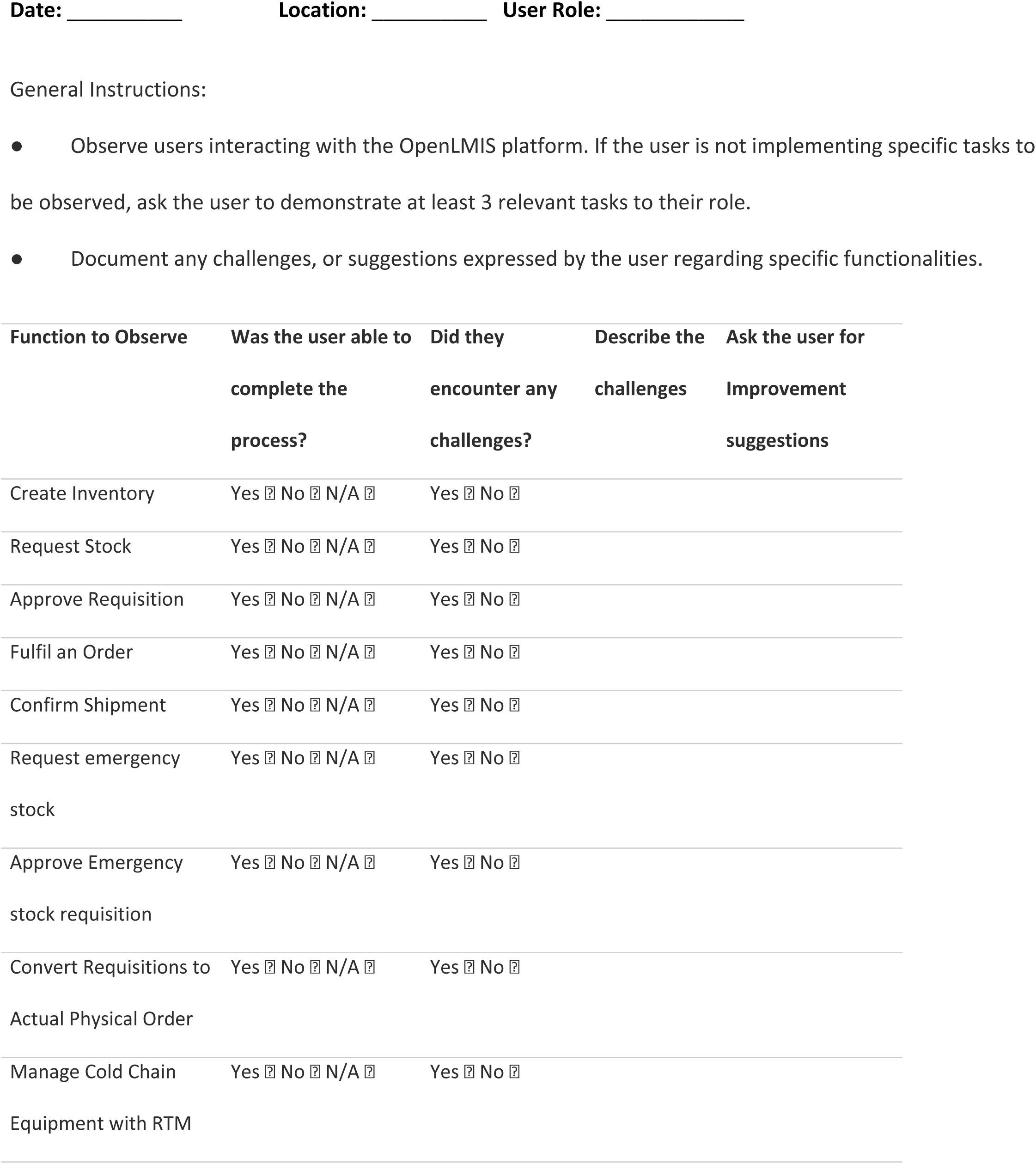
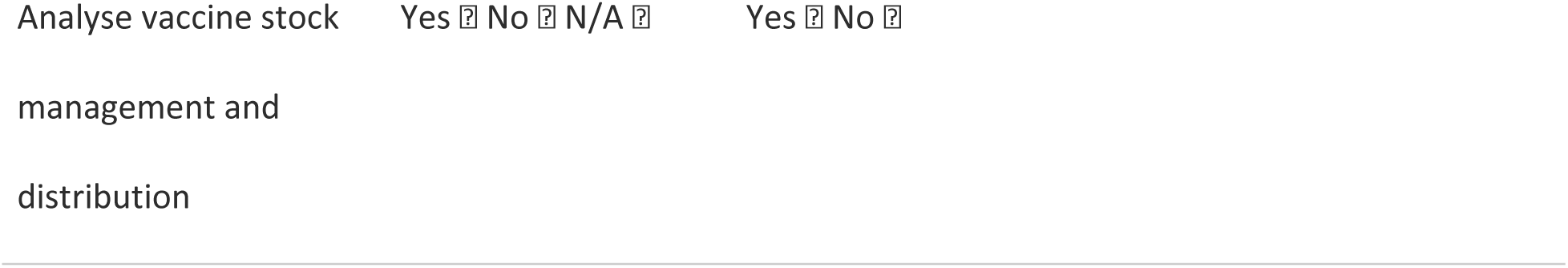

## References

1. Wasswa J, Oundo H, Oteba M, Ssozi J, Nakabugo K, Babiiha G, et al. Leveraging electronic logistics management information systems to enhance and optimize supply chain response during public health emergencies: lessons from COVID-19 response in Uganda. J Pharm Policy Pract. 2023;16(1):6. Available from: 10.1186/s40545-023-00517-4

2. Lydon P, Schreiber B, Gasca A, Dumolard L, Urfer D, Senouci K. Vaccine stockouts around the world: Are essential vaccines always available when needed? Vaccine. 2017;35(17):2121–6.

3. Gilbert SS, Bulula N, Yohana E, Thompson J, Beylerian E, Werner L, Shearer JC. The impact of an integrated electronic immunization registry and logistics management information system (EIR-eLMIS) on vaccine availability in three regions in Tanzania: A pre-post and time-series analysis. Vaccine. 2020;38(3):562–9.

4. Agarwal S, Perry HB, Long L, Labrique AB. Evidence on feasibility and effective use of mHealth strategies by frontline health workers in developing countries: Systematic review. Trop Med Int Health. 2015;20(8):1003–14.

5. Sarley D, Mahmud M, Idris J, Osunkiyesi M, Dibosa-Osadolor O, Okebukola P, et al. Transforming vaccines supply chains in Nigeria. Vaccine. 2017;35(17):2167–74.

6. Ugwu GO, Odii A, Bisi-Onyemaechi B, Ezema GU, Okeke A, Uzochukwu BSC, et al. Digital Technology Tool for Routine Immunization: Lessons Learned from Open Data Kit Intervention and Way Forward. Niger J Clin Pract. 2023;26 Suppl 1:S65–70. Available from: 10.4103/njcp.njcp_561_22

7. Gavi and The Global Fund. Qualified Software Solutions for Logistics Management Information Systems (LMIS): Country guidance on selecting LMIS. Gavi and The Global Fund; [cited 2025 Oct 14]. Available from: https://www.gavi.org/news/document-library/country-guidance-selecting-logistics-management-information-systems

8. OpenLMIS. How OpenLMIS in Nigeria helped save 850,000+ vaccine doses from expiry. OpenLMIS; 2025 Jun 4 [cited 2025 Oct 14]. Available from: https://openlmis.org/how-openlmis-in-nigeria-helped-save-850000-vaccine-doses-from-expiry/

9. Boeije H. Analysis in Qualitative Research. 1st ed. London: Sage Publications; 2010. 204 p.

10. Kaboré SS, Ngangue P, Soubeiga D, Barro A, Pilabré AH, Bationo N, et al. Barriers and facilitators for the sustainability of digital health interventions in low and middle-income countries: A systematic review. Front Digit Health. 2022;4:1014375.

11. Mantel C, Hugo C, Federici C, Sano N, Camara S, Rodriguez E, Al. Impact of electronic immunization registries and electronic logistics management information systems in four low-and middle-income countries: Guinea, Honduras, Rwanda, and Tanzania. Vaccine. 2025;54:127066.

12. Gebremedhin S, Shiferie F, Tsegaye D, Asefa W, Wondie T, Zeleke S, et al. Perspectives on the Performance of the Ethiopian Vaccine Supply Chain and Logistics System after the Last Mile Delivery Initiative: A Phenomenological Study. Am J Trop Med Hyg. 2024;110.

13. Sylla B, Ismaila O, Diallo G. 25 Years of Digital Health Toward Universal Health Coverage in Low- and Middle-Income Countries: Rapid Systematic Review. J Med Internet Res. 2025;27(1):e59042.

14. O’Brien N, Li E, Chaibva CN, Gomez Bravo R, Kovacevic L, Kwame Ayisi-Boateng N, et al. Strengths, Weaknesses, Opportunities, and Threats Analysis of the Use of Digital Health Technologies in Primary Health Care in the Sub-Saharan African Region: Qualitative Study. J Med Internet Res. 2023;25:e45224.

